# “I know that one day I will be cured, and all will be well”: Perspectives on acceptability of curative therapies for sickle cell disease in Tanzania

**DOI:** 10.1101/2025.07.31.25332489

**Authors:** Daima Bukini, Kassim Kassim, Collins Kanza, Aisha Rifai, Rahma Chanzi, Augostino Amando, Elianna Amin, Jonathan Spector, Julie Makani

## Abstract

While curative treatments for sickle cell disease (SCD) including hematopoietic stem cell transplant and gene therapies are largely restricted to high-resource settings, recent advancements have raised hopes that they will eventually become more accessible to patients in Africa, where the disease is particularly prominent and contributes to substantial morbidity and mortality. Understanding the factors that influence acceptability of curative therapies will therefore be important in developing strategies for successful adoption and implementation. To address this need, a qualitative study was conducted to gain insights into the perspectives of patients and families in Tanzania regarding curative therapies for SCD, focusing on factors affecting treatment acceptability. A total of 81 individuals participated through Focus Group Discussions or In-Depth Interviews. The results indicated substantial interest in current and prospective curative programs, with various factors influencing the attitudes of patients and caregivers differently. Study findings suggested, caregivers’ decisions were likely to be influenced by the burden of caregiving roles and the financial costs related to care for SCD. Patients were likely to accept the therapies if provided with information on safety of the therapies and requested for transparency of data from treated patients. In preparation for gene therapy trials or transplants, tailored engagement strategies for patients younger than 18 years old will be necessary as majority of SCD patients in Africa are children. Healthcare professionals must be well-informed to address questions and concerns from patients’ families. Future efforts should incorporate early involvement of patients and caregivers in Africa in programs related to curative therapies for SCD.

## INTRODUCTION

Until recently, hematopoietic stem cell transplantation (HSCT) was the only curative option available for individuals living with sickle cell disease (SCD) globally ^1,2^. While HSCT can provide rapid hematopoietic and immunologic reconstitution, the procedure is complex and is associated with risk of graft-versus-host disease and other complications, especially in patients without matched sibling donors^3^. Gene therapies (GT), through gene edition or addition, have since emerged as an innovative alternative and offer the potential to circumvent risks of HSCT^4^. Significant advancement have been since the first successful gene therapy trial for SCD was reported in 2017^5^. Approvals of gene therapy programs for SCD in the United States in 2023, and subsequently Europe provided hope to SCD stakeholders globally on the potential possibilities for safer curative options for SCD through gene therapies^6^.

Estimates indicate that more than 70% of 515,000 children born with SCD are in sub-Saharan Africa^7^. In Tanzania, the disease is of major public health concern, with up to 16,000 children born with SCD annually contributing to 6% of the overall under-fives mortality^8–11^. Recognizing this burden, the government of Tanzania has taken concerted steps to support curative therapy efforts for SCD^12^. In 2023, Benjamin Mkapa Hospital in Dodoma, the capital of Tanzania, performed the first HSCT for SCD in the country using related HLA-matched donors becoming one of the few centers in Africa offering BMT for SCD^13^. This achievement demonstrated that political will and infrastructural investment can lead to measurable successes in advanced therapy programs for SCD, laying groundwork for establishing gene therapy programs for SCD^14^.

Parallel efforts to champion curative interventions for SCD in the country are led by the Sickle Cell Programme based at Muhimbili University of Health and Allied Sciences in Dar es Salaam. The program has been focusing on establishing HLA donor registry to facilitate bone marrow transplant and establishing partnerships aiming to facilitate implementation of gene therapy trials for SCD in Africa^15^. Of notable achievement is the work implemented through partnerships with Novartis Biomedical Research (NIBR) to develop the consensus-guided Target Product Profile (TPP) for both invivo and exvivo gene therapy products for SCD, with potential application in Africa^16^.

In addition, the program implemented comprehensive patient engagement strategies to gather knowledge and understanding related to curative therapies from patients and caregivers. In high-income countries such as the United States (US), several studies have explored factors influencing the acceptability of curative therapies for SCD, providing valuable insights into patient and caregiver attitudes toward HSCT and gene therapies^17,18^. Other studies demonstrated that factors such as socioeconomic status, cultural beliefs, and access to information significantly influence decision making for patients and caregivers in embracing new therapies^19^. However, these studies have primarily focused on populations where access to healthcare and socioeconomic contexts differ significantly from those in sub-Saharan Africa. Understanding factors that influence the acceptability of curative therapies for SCD, is expected to help inform effective adoption and implementation strategies at community level.

This paper reports a phenomenology-based qualitative study exploring the perspectives of patients and caregivers on HSCT and gene therapies, specifically focusing on factors influencing the acceptability of curative therapies for SCD.

## METHODS

A phenomenology-based qualitative study was designed to understand patients and families’ perspectives on curative therapies for SCD, defined in this project as HSCT and GT. Phenomenology is a research approach that evaluates individuals’ lived experiences to understand their thoughts and feelings about a specific topic^20^. The methodology, which took place through Focus Group Discussions (FGDs) and In-Depth Interviews (IDIs), was selected to enable researchers to discover a nuanced view of how curative therapies would impact study participants’ lives. The study was conducted after a series of Deliberative Engagement (DE) sessions with the participants to support their health literacy relating to curative therapies for SCD and to facilitate successful translation of study materials into Kiswahili. DEs have been previously used to engage with participants in Africa on the topics of human genome editing and return of genetic findings ^21,22^.

**Figure 1:**
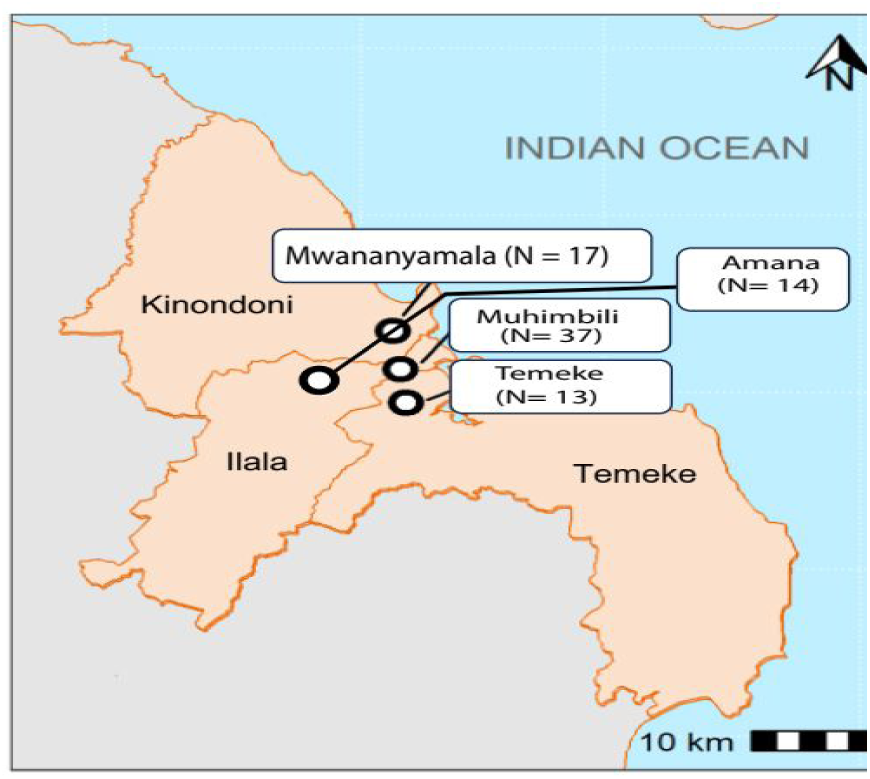
Map of Dar es salaam, showing the four major SCD clinics and number of individuals recruited for this study from the clinics study (patients and caregivers)

**Figure 2:**
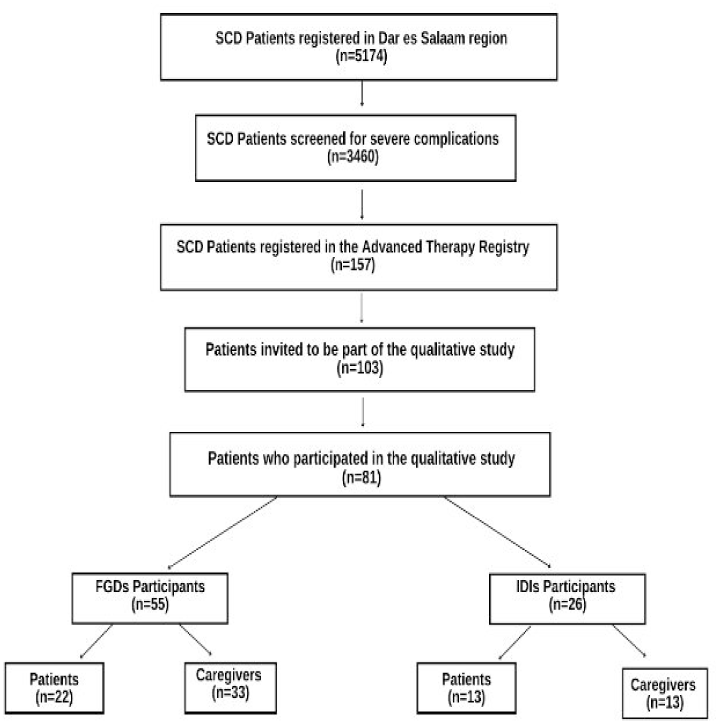
Recruitment flow chart for study participants included in the study showing categories for both focus group discussions and in-depth interviews.

### Study setting and recruitment strategies

This study was conducted in the Dar es Salaam region of Tanzania. As of December 2024, a total of 5,174 individuals with SCD were registered through four regional referral hospitals (Amana, Temeke, Mwananyamala and Muhimbili) that have dedicated weekly SCD clinics for pediatric and adult patients. More than 80% of registered patients are below 18 years of age. A subset of 157 are included in the Advanced Therapy Registry, a dataset of individuals that are more severely impacted by complications of SCD and who are considered priority preferred candidates for curative therapies. Amongst which, the research team invited 103 individuals were randomly invited to be part of the study. Eighty-one (81) individuals agreed to participate, out of which 45 were caregivers and the remaining 36 participants were patients.

### Demographic characteristics of the study participants

The demographic profile of study participants is summarized in Table 1. There were 54 females (67%) and 27 males (33%). The majority were aged 21-30 years (42%), followed by 31-40 years (32%), and smaller proportions in other age groups. Most participants were engaged in small businesses (49%), while others worked from home (22%), were retired (9%), were students (7%), or were formally employed (1%). Forty-two percent of participants completed primary-level education, 37% completed secondary-level education, and fewer attained higher education levels (6% and 4% completed university and college training, respectively). Most participants reported having one child with SCD (27%), with smaller proportions (14%) reporting two or more children.

**Table 1:**
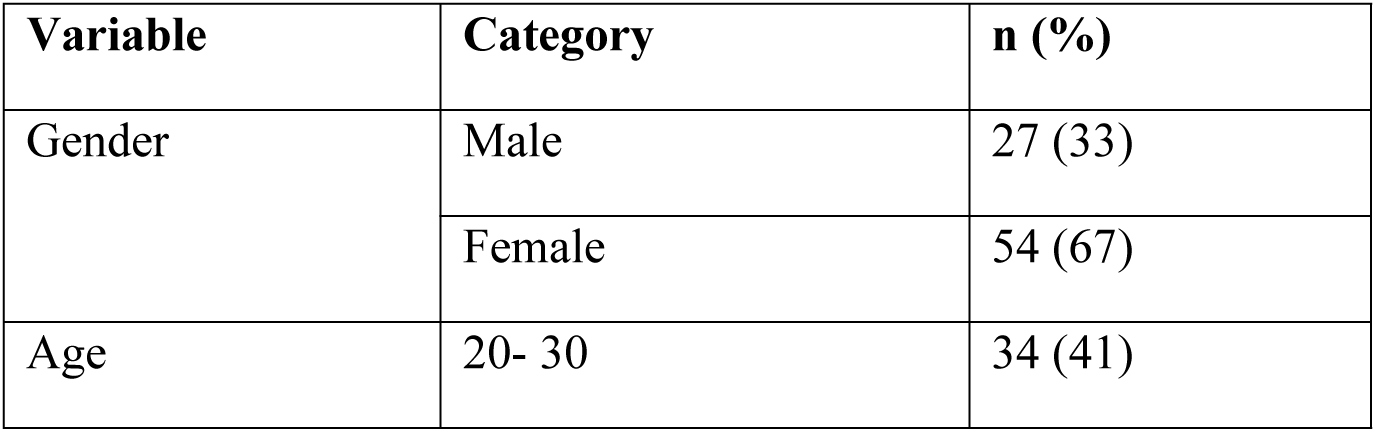
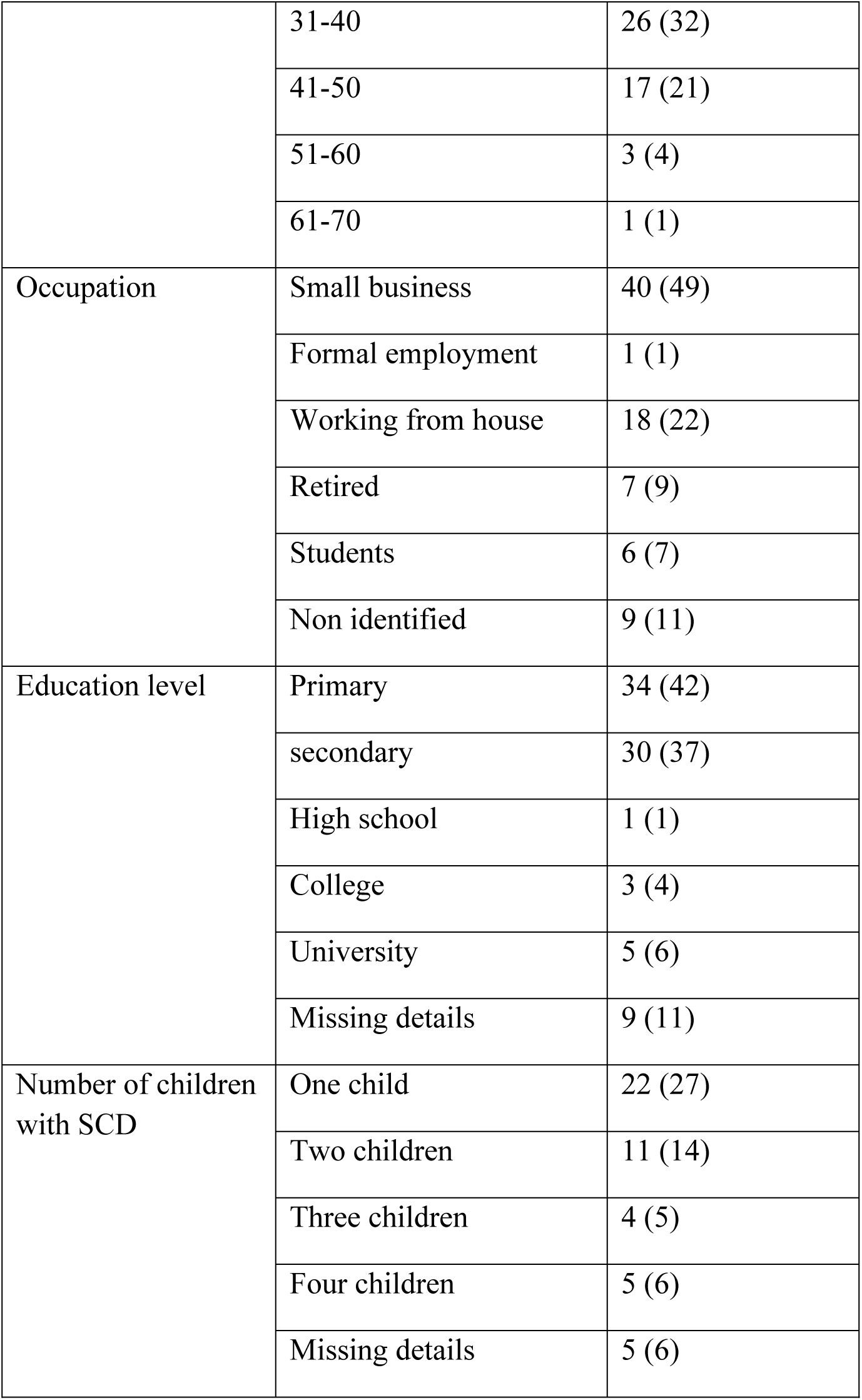
Demographic characteristics of study participants.

| Variable | Category | n (%) |
| --- | --- | --- |
| Gender | Male | 27 (33) |
|  | Female | 54 (67) |
| Age | 20- 30 | 34 (41) |
|  | 31-40 | 26 (32) |
|  | 41-50 | 17 (21) |
|  | 51-60 | 3 (4) |
|  | 61-70 | 1 (1) |
| Occupation | Small business | 40 (49) |
|  | Formal employment | 1 (1) |
|  | Working from house | 18 (22) |
|  | Retired | 7 (9) |
|  | Students | 6 (7) |
|  | Non identified | 9 (11) |
| Education level | Primary | 34 (42) |
|  | secondary | 30 (37) |
|  | High school | 1 (1) |
|  | College | 3 (4) |
|  | University | 5 (6) |
|  | Missing details | 9 (11) |
| Number of children with SCD | One child | 22 (27) |
|  | Two children | 11 (14) |
|  | Three children | 4 (5) |
|  | Four children | 5 (6) |
|  | Missing details | 5 (6) |

### Development of data collection tools

The process of developing data collection tools started during DE sessions with prospective participants. Five DE sessions were attended by an average of 50 participants per session, including both patients and caregivers. The first three DE sessions, conducted between March and June of 2021, included clinicians from local health centers and focused on understanding baseline levels of health literacy among participants regarding HSCT and GT. Two additional sessions, held in June 2022 and April 2023, were structured to enable focused discussion in key areas that were identified in the initial sessions to require further deliberation with researchers and clinicians. Knowledge gained through the DE sessions were used to inform the development of FGD and IDI guides for patients and caregivers. The DE sessions supported development of appropriate Kiswahili terminologies and translation that would be readily understood by participants during the study procedures. The DE sessions were also used to purposefully identify participants to be invited to participate in the study, with a particular focus to those who provided insights requiring in-depth interviews or discussions. FGD and IDI guides were pre-tested through one FGD involving eight patients and two IDIs with caregivers to evaluate the format of the guides and language comprehension. Box 1 summarizes key topics that were included in the FGDs and IDIs with patients and caregivers. Snacks and refreshments were offered to participants and accompanying children during FGDs and IDIs.

#### Box 1 Key topics in Focus Group Discussion and In-Depth Interview guides for patients and caregivers derived from Deliberative Engagement sessions

Tell me what you have heard from different people about HSCT and GT. Prompts: Given what you’ve heard, what do you think about it? What do you think you would do?

What did you think initially about HSCT and GT, and how did these ideas change over time?

If HSCT or GT was available for you or your child, how would the decision-making process between the two therapies be the same or different?

HSCT requires a caregiver to be in the hospital for approximately three months and come monthly for an additional six months for follow up. Tell us how the family will negotiate the different roles and obligations in the households and hospitals. Prompts: Caregiving roles at homes and hospitals; how financial obligations will be managed in the families, food, school fees; how are men and women impacted differently during the decision making.

Tell me about the best ways to get information about HSCT and GT to families affected by SCD, including patients and their caregivers. Prompts: pamphlets, workshops, art exhibitions, online.

What information do you believe is important to know regarding HSCT and GT?

What do you think about the approximate cost of HSCT in Tanzania?

What would the ideal pricing model for HSCT or GT be? Prompts: Private, out-of-pocket, insurance-based, sponsorship (public or foreign). Why?

What would the ideal situation look like for you when being treated with more advanced technologies in terms of cost, location, and service aspects?

### Data Collection Methods

#### a) Focus Group Discussions

Eight FGDs were conducted with 55 participants; four FGDs involved patients and four FGDs were focused on caregivers. Each FGD included at least five participants with the largest group having a total of nine participants. Sessions were moderated by two trained research personnel (KK, CK) and one observer (AR, RC or AA) and lasted approximately 1 to 1.5 hours. A total of 23 patients and 32 caregivers participated in FGDs. Most caregivers were mothers of children with SCD (n=24), and four men participated. For FGDs involving patients, nine men and 14 women participated.

**Table 2:**
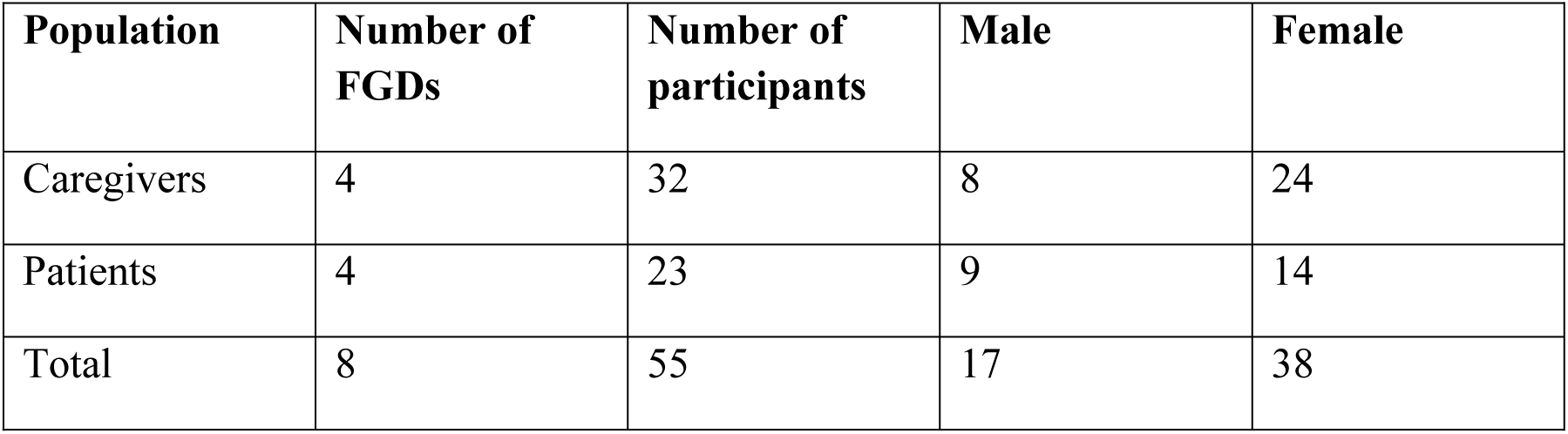
Details of Focus Group Discussions.

| Population | Number of FGDs | Number of participants | Male | Female |
| --- | --- | --- | --- | --- |
| Caregivers | 4 | 32 | 8 | 24 |
| Patients | 4 | 23 | 9 | 14 |
| Total | 8 | 55 | 17 | 38 |

#### b) In-Depth Interviews

Twenty-six IDIs were conducted with patients (n= 13) and caregivers (n= 13) to gain a deeper understanding of individual perspectives on curative therapies. For caregivers, one participant was a father and the remaining 12 were mothers of children with SCD. Of the 13 patients who participated, 6 were male and 7 were female. Interviews were conducted for up to 45 minutes on a one-on-one basis in a private conference room at the MUHAS Sickle Cell Center (interviews performed by DB, KK or CK with AI, RC or AA observing).

**Table 3:** Details of In-Depth Interviews.

| Population | Number of IDIs | Number of participants | Male | Female |
| --- | --- | --- | --- | --- |
| Caregivers | 13 | 13 | 1 | 12 |
| Patients | 13 | 13 | 6 | 7 |
| Total | 26 | 26 | 7 | 19 |

### Data analysis

Thematic analyses were conducted on outputs from FGDs and IDIs according to standard qualitative methods. Research team members (DB, KK, CK, AR, RC, and AG) reviewed transcriptions from digital recordings and handwritten notes. Initial coding was done individually, followed by weekly meetings over several months to review and reach consensus on the codes. A codebook was developed using NVivo (version 14) for coding (process led by DB and KK). The team identified potential limitations in question phrasing and revised some questions to enhance insights. Consensus on data patterns led to grouping findings into themes.

## RESULTS

The thematic analysis of FGDs and IDIs with patients and caregivers identified several themes affecting the acceptability of curative therapies for SCD. While most participants were open to exploring a cure, certain factors influenced their decisions, as shown in the diagram and summary below.

**Figure 3:**
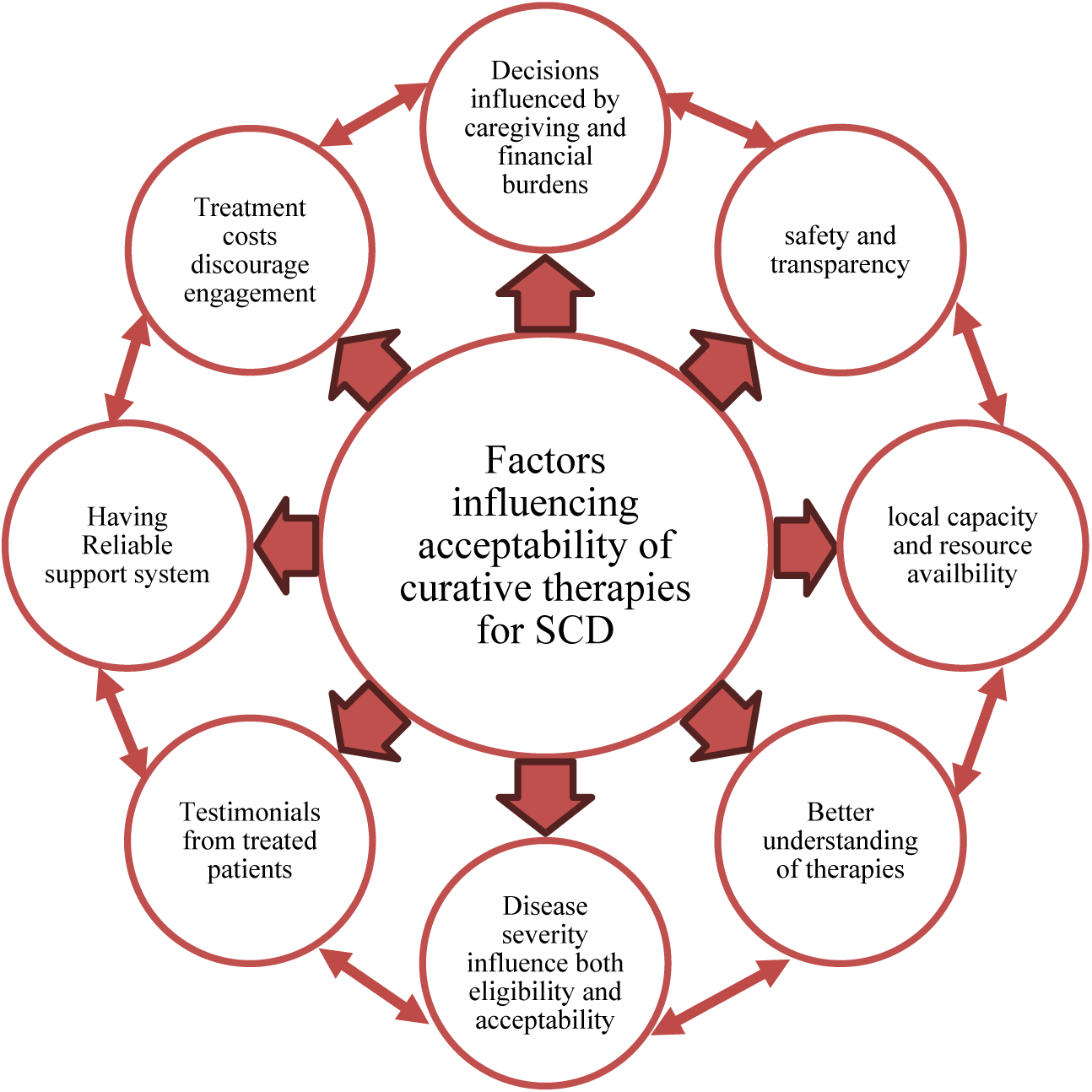
Schematic representation of major themes identified in the analysis influencing acceptability of curative therapies for SCD from patient and caregiver perspectives.

### Motivation for one-time cure solution linked to caregiving challenges

The majority of participants, particularly caregivers, indicated a willingness to consider either Hematopoietic Stem Cell Transplantation (HSCT) or Gene Therapy (GT) as a potential “one-time”cure for the disease. For caregivers, this motivation was primarily driven by the significant burden of caring for a child or children with Sickle Cell Disease (SCD), which is further exacerbated by additional caregiving responsibilities within families, financial constraints, and general challenges in accessing healthcare. Caregivers believed that curative therapies could potentially result in substantial improvements in the quality of life for both their children and themselves. For example, one caregiver stated, *“When I first received this information [about a potential cure for SCD], I was truly relieved and felt that now my children have found a savior. We parents face very tough challenges, and it can be difficult because of our financial situation. Sometimes a child gets sick in the middle of the night and you have nothing in your pocket, yet you need to take them to the hospital. So, we go through many challenges” (Caregiver, FGD, July 2023)*.

### Motivation influenced by safety of therapies

Participants’ willingness to undergo curative therapies was significantly influenced by assurances regarding the safety of these treatments and transparency about their potential side effects. This perception was more evident in interactions with patients compared to caregivers. While many generally embraced the potential for curing Sickle Cell Disease (SCD) and the possibility of being pain-free, some remained concerned that unknown risks could worsen their condition or introduce new health issues. A quote below from the patient who participated in FGD number 3, compared the new treatments to when Hydroxyurea was initiated, and the concerns that existed on the side effects. *“So, even when hydroxyurea arrived, the questions were the same, like, what is this thing, how does it work? We were worried, but then the questions were answered, and we got used to it*.

*We are hoping the same will happen with these new treatments; the important thing, is if they (treatments) are safe. We don’t want to come out with changed skin color or other side effects; they just need to be safe and affordable. (Patient, FGD, January, 2024)*

### Perspectives on local capacity for administration of therapies and access considerations

Participants focused on the availability of HSCT services in Tanzania. Some were unsure if these services could be provided locally. Concerns included the resources and expertise needed to support them adequately, as exemplified by the caregiver’s statement, “First*, I was concerned that we might have limited expertise to carry out such significant tasks. Cost was not a big deal because anything is possible if it has been done elsewhere, and we have the greatest burden of patients. I think it’s possible, but my biggest concern was the professionals because as we see, there are many services available elsewhere that we should also have, but they have taken too long to arrive. Even if they were to arrive, some are not accessible [inaudible]. However, for now, we have seen things progressing. What remains is to say that we should fight for these services to reach our patients”(Caregiver, IDI, July 2023)*.

The HSCT program in Tanzania is currently eligible for children below 12 years of age with matched sibling donors. Some patients in the study were not motivated to follow up further on the service because they are not eligible to receive the therapies. For example, one participant was curious about the HSCT services they learned about in local media only to find out that they were not eligible: “*The first day when I realized, I asked, is this really possible? But as I continued following up, I saw that it is indeed possible. Later, I saw Millard Ayo had posted that a child was in parliament and had received a bone marrow transplantation, I don’t know if it was his sister, but [not clearly audible], then it became a cost issue, and then the statement that it was under 5 years old, I said I’m not involved there, I didn’t pursue it, I left it alone because even if I followed it up, I’m not involved, I’m not in that group. I asked myself, “Why is it possible for them and not for us?”(Patient, IDI, October 2023)*.

### Motivation enhanced with improved understanding of curative therapies

Patients and caregivers who received comprehensive information prior to interactions gained sufficient understanding of curative therapies, leading to increased openness to the possibility of a cure for Sickle Cell Disease (SCD). Caregivers expressed interest primarily in the potential benefits and access issues related to the therapies, while patients were more concerned with the treatment processes and potential side effects. One patient specifically requested additional clarity on the procedures, including details about the cost, duration of the treatment, and the expected length of effectiveness: “*I think my concern comes back to the fact that I still don’t have enough education. I keep asking myself questions, as I told you. If I make this change, what are the benefits and the drawbacks? That’s why I continue to worry. After making this change, will I be a frequent hospital visitor? Or will it reduce my condition somewhat? For example, as I told you, my hip was deteriorating, and they had to work hard to fix it. I have seen success when they replaced the hip, but I still wonder about these new therapies [reference to exchange transfusion, HSCT, and GT]” (Patient, IDI, October 2023)*.

### Disease severity as a motivation to explore curative therapies

Patients and families with severe disease complications were more receptive to therapies than those with mild cases. The potential for a cure or significant improvement outweighed the uncertainties or risks. In a FGD with patients, disease severity was seen as crucial for accessing therapies, with frequent care or hospitalizations indicating a greater need. One patient stated: “*However, what might motivate a patient to undergo treatment is their health condition, which could increase the need for the procedure. For instance, some patients can’t go more than two months without being hospitalized, which creates a greater need for this treatment” (Patient, FGD, October 2023)*

Similarly, caregivers of children with severe complications often associate the need for treatment with their distress at witnessing their children’s suffering and being unable to alleviate it. One caregiver stated: “*I felt relieved because when you live with someone and see how much they suffer, it pains you even more. As a parent, seeing the pain my child endures makes me feel terrible. So, when I heard this news, I felt very relieved.” (Caregiver, IDI, January 2024)*

### Testimonials from patients who have been cured of SCD

Patients and caregivers often rely on success stories from those cured by new treatments to guide their decisions. These stories significantly impact their choices, with most following social media updates (e.g., WhatsApp groups) for news on SCD cures. Such stories increase their curiosity and raise questions about eligibility and access, as demonstrated by one caregiver’s statement: “*I would like to know about the children who were announced to have undergone bone marrow transplantation. How is their progress so far? How are they inspiring us?” (Caregiver, FGD, October 2023)*

### High costs of treatments discouraging interests for cure

In comparison to discussions on access to comprehensive care services, there was limited interest in engaging in discussions about curative treatments due to the associated costs. This was particularly relevant for families who could not afford current care and would therefore direct the conversations towards questions on access to existing therapies such as Hydroxyurea.

Enthusiasm for curative options tends to diminish when the focus shifts to the costs of treatment. One caregiver stated: “*But what really discouraged me was the cost. Considering I am on my own, I wondered how I would afford it. The fact that I am struggling to access this service is troubling me; I don’t see how my child could reach this point. That’s when I realized it would be impossible.” (Caregiver, FGD, October 2023)*

### Motivation for cure linked to presence of family support during and after treatment

Some patients’ willingness to receive therapies depended on family support. This was crucial because treatments required long hospital stays and post-hospitalization financial assistance. As shown in a quote from one of the FGDs, patients wanted their families and friends involved for support during and after treatment: “*First, let’s work together with relatives or friends to prepare your environment because after undergoing treatment, you will be dependent for a certain period. Therefore, until you recover, you need to be under close care. Thus, working with relatives and friends to create a supportive environment can allow one to recover quickly, and some can provide love and affection which can speed up the recovery process and allow one to return to their duties.” (Patient, FGD, July 2023)*

### Religious and cultural considerations did not impact interest in curative therapies

During interviews and discussions with patients and families, one of the key areas explored was the potential influence of religious or socio-cultural beliefs on acceptability and decision-making processes. The FGD and IDI guides contained specific questions to examine this relationship. However, the data did not indicate any direct influence of religious or socio-cultural beliefs on the acceptability of SCD cure among families or patients. Both caregivers and patients indicated that procedures like transplants, which aim to cure a child from disease, align with the intention of alleviating suffering and do not conflict with traditional cultural norms. Additionally, religion was not seen as a barrier, as it supports seeking treatment for health issues. Both cultural and religious factors were perceived as supportive in the decision-making process. One patient stated: “*Regarding cultural practices, there are no issues. I mean, when it comes to cultural practices, there are no problems because I believe that procedures like transplants and similar things aim at the intended purpose and do not bring anything different. So, with cultural practices, there are no issues. And with religion, there are no issues either because you are supposed to seek treatment to solve your problems.” (Patient, IDI, April 2023)*

There was, however, one comment from a patient during an FGD who wondered aloud about the appropriateness of curing SCD on religious grounds: ***“****I have never sat down with religious leaders, but in faith, they tell you that disease is from God or that is how God created the person, so you cannot change a person’s disability or restore them to a normal state.” (Patient, FGD, January 2024)*.

## DISCUSSION

This study aims to gather perspectives from caregivers and patients to understand issues related to the acceptability of curative therapies for SCD in Tanzania. There is significant interest among both patients and caregivers involved to benefit from current and prospective curative programs for SCD. In both FGDs and IDIs, participants provided detailed perspectives on what factors may influence their intention to seek for cure.

On the topic of eligibility, both patients and caregivers agreed that severity of the disease should be the top eligibility indicator, which was also considered important in other similar studies^23^. Suggesting that patients presenting with severe complications of the disease will more likely be willing to opt for cure compared to others. Although, in some instances patients who have severely suffered from the SCD complications may be in-eligible to receive the treatments to avoid exacerbation of their condition or poor clinical outcomes^24^. Aligning understanding of the patients and families on the indications for transplant or potentially gene therapy will help balancing their expectations and weighing on the risks-benefit of the treatments.

High treatment costs were a huge deciding factor on acceptability of the therapies for both patients and families. While the current transplant program at BMH is still funded through the government but the costs of the treatments are estimated at 50M Tanzanian shillings (∼ 20k USD), which still is not affordable to most families. Engagement with the public and private insurance schemes to devise cost sharing mechanisms to reduce potential financial burden from families will allow more eligible patients to access the services. The current cost for GT (∼ 2MUSD) is out of reach for SCD patients in Africa, who stand to benefit from the treatment^6,25^. In the US, as an effort to increase access Bluebird Bio and Vertex Pharmaceuticals will partner with the the cell and Gene Therapy (CGT) Access model to lower the costs of the treatments^26^.

This initiative builds on the anticipation for countries in Africa with high burden of SCD to explore access negotiation with the pharmaceutical companies to lower down the costs of the treatments. Other efforts to lower costs for GT for patients in LMICs are led by Global Gene Therapy Initiatives^27^.

Reliable family support was considered essential in deciding to participate because of the support required during and after transplants. Involvement of the family members at the beginning to understand the importance of their supportive roles pre- and post-transplant is considered important throughout the treatment. Caregiving for patients with SCD in the settings is largely the work involving extended network of care^28^, and therefore their participation as caregivers is necessary for patients or families to decide on opting for curative therapies, that more often require long treatment plans. This necessitates comprehensive discussions with patients and families on the required social and clinical support to decide on the right timing for the transplant and who will provide primary care. For adult patients with families, support is needed not only during treatment but also for children left behind while the parent undergoes treatment. As mothers and women in the settings are particularly more responsible for the burden of care of the children with SCD^29^, it is essential to carefully analyze the level of work and sacrifices required or imposed to the mothers during treatment to avoid exacerbating their burden of care. Hospital based programs linked to community health workers or nurses to support mothers pre- and post-transplant could help ease their responsibilities. The requirement of reliable support also needs to be carefully reviewed to avoid favoring patients from well-off families, who are capable of providing supportive care required pre- and post-transplants.

Adult patients (above 18 years of age) included in the study were considered capable of comprehending information about the therapies, asking questions, and seeking clarity on areas requiring additional details. Specific interests were on the safety of the therapies and potential side effects. Therefore, it is important for initial information sessions with the patients to focus on understanding the details and processes of the therapies and how they might affect the patient’s quality of life during and after treatment. Presenting the information in a comprehensible format detailing potential safety issues will be useful. Designing tools that visualize the information through pictorial formats or video recordings could help support comprehension^30^.

On the decision making, as majority of the patients are children below 18 years of age, decisions are potentially being made by the parents or caregivers. And therefore, ethically important to ensure that decisions are made on the best interests of the child after thorough assessment of the motivation for treatment, providing adequate information on both the potential benefits and risks of the treatments to support the decision-making process. In some instances, the caregiver’s motivation for cure were linked to the caregiving and the financial burdens. Assessment of the motivation factors at the initial stages will be needed to protect the interest of the children from unnecessary risks of treatments that may not be of benefits. Devising mechanisms to involve children (12-17 years) in discussions during the sessions will help bridge the information gap between children and caregivers and assess their willingness to participate^31^. Simple pictorial representation to support engagement of children across different age groups to facilitating assent may be necessary in this situation.

Testimonials from cured patients, including data on clinical progress and adverse events related to the treatments, should also be transparently available to patients and families. This was considered crucial by patients to support their decision to opt for curative treatment, finding consistent with similar studies^23^. Victoria Gray testimonial as the first patient cured of SCD through gene therapy has been powerful in adding the reality of potential cure to many SCD patients globally^32^. Jimi Olaghere’s recent voyage to Tanzania reaching the summit of Mount Kilimanjaro was also an equally powerful testimonials of the possibility of limitless potential after being cured of SCD^33^. As more testimonials and clinical data are widely accessible to patients will support in making informed decisions on what are the best treatment choices as they continue to explore different paths to cure.

Patients and families remain concerned about challenges in accessing basic comprehensive care. Researchers and clinicians must address questions about why investing in high-cost treatments will address the overall burden of SCD in the region. Some participants questioned infrastructural preparedness and local capacity to support curative therapies. Questions include who will perform the procedures, their training, and where the program will be conducted. These concerns could be addressed by having clear referral pathways to curative therapies and integrating the programs within current comprehensive care services. A continuum of SCD care from lower healthcare facilities to tertiary facilities offering HSCT will need to be established and aligned within SCD care provision. Healthcare professionals will also need to be well-informed to address and clarify questions and concerns from families. Looking ahead in Tanzania, the referral system that will be built may be leveraged as a pathway for GT^15^.

## CONCLUSION

Although relatively few centers currently provide HSCT to patients with SCD in Africa, and there are no approved gene therapies in the region, understanding factors that may influence potential uptake is important to align future services with the expectations of key stakeholders. The findings from this study underscore vital topics that necessitate engagement at various levels with patients and caregivers. Addressing these concerns through targeted engagement and informed strategies will help to maximize a patient-centered approach that aligns medical advancements with the unique needs and expectations of the affected population. This proactive engagement is key to fostering trust, improving healthcare outcomes, and ultimately enhancing the quality of life for patients with SCD in Africa.

## ETHICAL CONSIDERATIONS

The study was approved by the Muhimbili University of Health and Allied Sciences Research Ethics Committee. Written consent was sought from the participants The study involved research participants who are 18 years and older to ensure that all participants have reached the legal age to consent.

## CONFLIC OF INTEREST

EA are JS are employees of Novartis Biomedical Research

## Data Availability

All data produced in the present work are contained in the manuscript.

## ACKNOWLEDGEMENTS

We thank parents and patients who have participated in this research.

